# Intraluminal pressure in central venous hemodialysis catheters during power injection of contrast media

**DOI:** 10.1101/2024.07.23.24310903

**Authors:** Nicholas A White, Aart J van der Molen, Ronald W A L Limpens, Jacinta J Maas, Koen E A van der Bogt, Tim Horeman, Joris I Rotmans

**Affiliations:** Department of BioMechanical Engineering, Delft University of Technology, Delft, The Netherlands; Department of Internal Medicine, Leiden University Medical Centre, Leiden, The Netherlands; Department of Radiology, Leiden University Medical Centre, Leiden, The Netherlands; Department of Cell and Chemical Biology, Leiden University Medical Center, The Netherlands; Department of Intensive Care, Leiden University Medical Center, The Netherlands; Department of Surgery, Leiden University Medical Center, The Netherlands; University Vascular Centre West, Leiden | The Hague | Delft, The Netherlands

## Abstract

**Background:** Central venous catheters (CVCs) provide direct access to the central circulatory system, commonly used in hemodialysis and intensive care units for drug administration. Although uncertified for the procedure, CVCs are sometimes used for power injection of contrast medium (CM) during CT scans to avoid peripheral intravenous catheter placement. Previous studies suggest this practice is safe, but incidents have been reported. This study aims to measure intraluminal pressure during CM injection through CVCs and assess its impact on the luminal surface to guide responsible clinical use.

**Methods:** Strain gauges were applied to the exterior walls of four samples from three different types of unused CVCs. These gauges measured material deformation due to intraluminal pressure during CM injections at rates of 4.5 mL/s and 8 mL/s, each performed five times. Strain data were calibrated against known pressures in a static system. The CVCs were then either pressurized until bursting or subjected to microscopic analysis of their luminal surfaces.

**Results:** Intraluminal pressures measured (97-545 kPa or 14-79 PSI) were below the burst pressure (779-1248 Kpa or 113-181 PSI) in all instances. Strain regression analysis shows a statistically significant (p<0.05) trend over 10 injections in almost all CVCs tested, indicating material fatigue. Surface microscopy revealed surface micro-cracks from repeated injections, suggesting material damage.

**Conclusions:** The intraluminal pressures from power injections of CM are sufficiently low to prevent CVC bursting. While incidental use for CM injection appears safe, repeated use may cause material damage

## Introduction

Hemodialysis patients frequently utilize a central venous catheter (CVC) as vascular access to transport blood to and from the dialysis. Similar CVCs may also be used for administration of drugs to patients in intensive care units. For diagnostic purposes, these patients may need computed tomography (CT) scan with radiopaque contrast medium (CM). Bolus intravenous injection of CM using a power (pressure) injector is the preferred method for CM administration for CT examinations of the neck, chest, and abdomen.^1, 2^ This is usually achieved by placing an intravenous needle into a peripheral vein in the forearm. However, hemodialysis patients often have poor peripheral veins, necessitating preservation for future arteriovenous access surgery. Despite studies indicating safe pressure levels for CVCs, even non-hemodialysis CVCs with smaller diameters,^3–9^ concerns remain about catheter-related complications due to high pressures, with several incidents of rupture and dislocation reported.^10–12^

CVCs are regulated under the European Medical Device Regulation in the EU,^13^ and similar bodies elsewhere, like the FDA in the United States.^14^ Manufacturers of medical devices only receive market approval for their products when the relevant requirements for performance and safety have been met.^13^ These requirements relate only to the intended use of the device, as specified by the manufacturer. Expanding use cases significantly increases the evidence required for approval, leading manufacturers to limit intended uses due to associated costs. This restriction is detailed in the device’s Instructions for Use, and off-label use shifts liability from the manufacturer to the healthcare professional.

Most standard CVCs are not certified for CM injection^15^ which can cause hesitance in administering CM through these devices. While successful use has been reported widespread,^15–17^ the high viscosity of CM causes an intraluminal pressure that is higher than, e.g., saline or blood, especially at high flow-rates. Manufacturers can claim that the CVCs are not rated for this elevated pressure as it may cause damage to the luminal surface of the CVC. While typical CM flow velocities are around 3-6 mL/s,^18–20^ in vitro measurements showed rupture does not occur even with much higher flows.^5^ In clinical use, pressures in the power injector are limited to maximum of 2000-2250 Kpa (300-325 PSI). Clinical guidelines for CM injection through CVCs are rare.^21^

Although power injectors can usually record the pressure applied to administer CM, it cannot be claimed that this pressure is constant throughout a dynamic, flowing system. Indeed, there must be a pressure gradient for fluids to flow from the power injector to the venous circulation of the patient. The difference in pressure depends on many variables, including viscosity, flow velocity, friction of the tube surface, and geometry. While this means the pump pressure overestimates the pressure downstream, (e.g., in a CVC), the non-uniform geometry of CVCs could cause local increases in pressure and wall stress. It is crucial that pressure is measured at the correct location to obtain the most accurate data on intraluminal pressure in CVCs when used for CM injection to properly assess safety.

Pressure transducers in a flow circuit interfere with the flow, often requiring a three-way connection that introduces static fluid dead space. This dead space can impede accurate pressure measurement due to its lack of direct pressure response to fluid flow and the flow distortions it introduces. In systems with rapidly changing flow rates, dead space can dampen fluid oscillations and distort pressure readings, contributing to significant pressure measurement differences in previous studies (14-483 kPa (2-70 PSI) at 4.5 mL/s with similar catheters). ^3, 4, 7, 9^

As a non-invasive method to measure pressure without the intrinsic drawbacks of pressure transducers, strain gauges may pose a solution. These contain thin pieces of wire that can be stuck onto a surface that measure deformation, or strain, which can be directly related to intraluminal pressure. Additionally, strain gauges are typically used to determine material wear and fatigue over time. As such, they may provide information relating to such effects occurring in the CVC material with CM injection.

The present study aims to equip healthcare professionals with the knowledge required to make informed decisions regarding the usage of commonly used CVCs for power injection of CM by presenting a comprehensive study of intraluminal pressure during injection, material fatigue, bursting pressures, and surface analysis.

## Materials and methods

### Experimental setup

The CVCs are tested by repeated injection of CM and recording the response with the strain gauge. The strain gauges are then calibrated with a pressure transducer in a closed system with static pressure in which pressure is equal everywhere and in all directions. Calibration is performed after the CM injection; the pressure necessary to properly calibrate the sensors must exceed the pressure induced by the CM, and therefore may damage the CVCs and distort the pressure recordings if performed prior to the power injections. Finally, the CVCs are either burst or dissected for microscopic surface analysis. The experimental setup is displayed in Figure 1.

**Figure 1:**
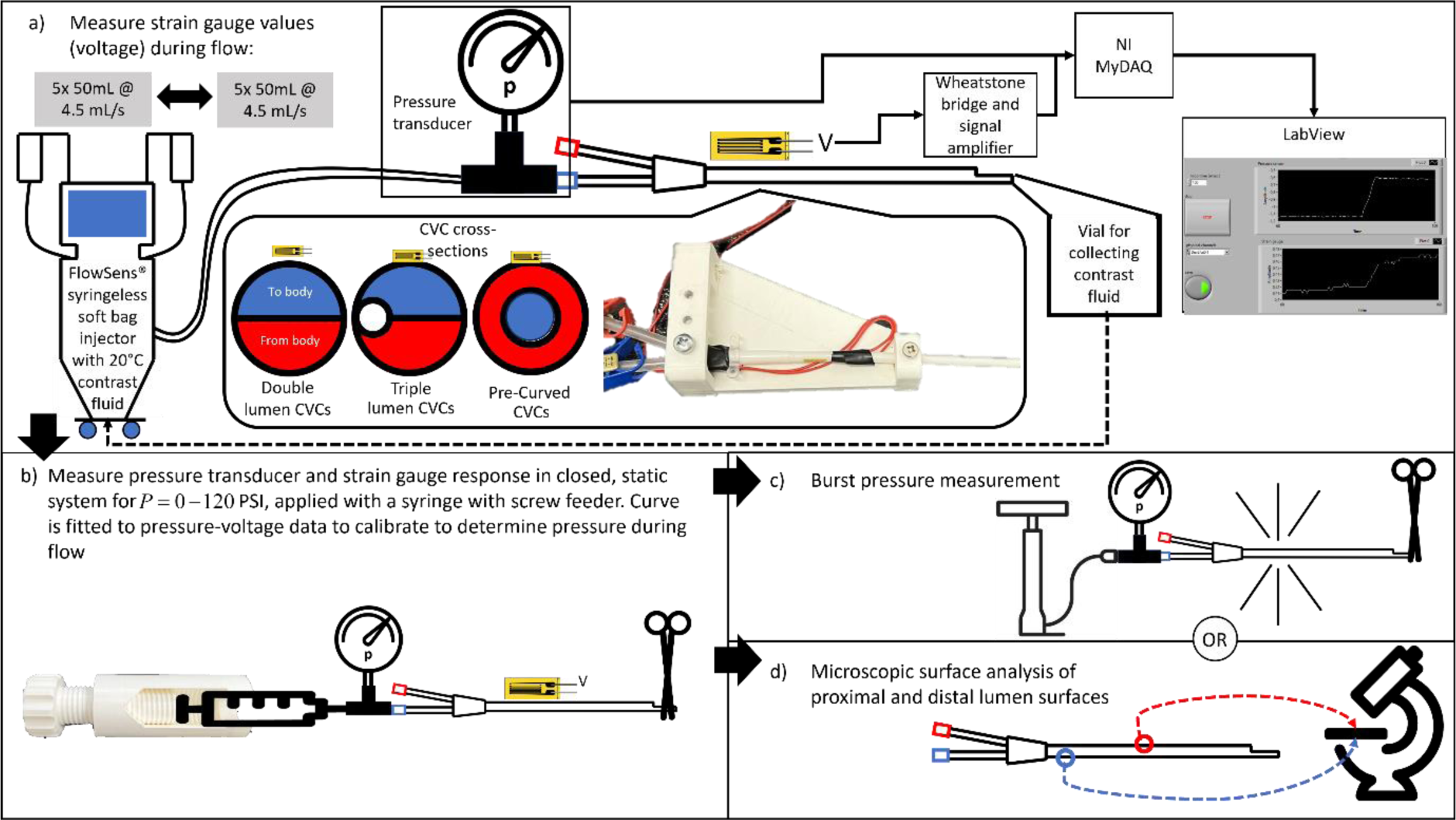
The experimental setup for testing the central venous catheters: a) the catheters with strain gauges are clamped and connected to a contrast medium injector with a pressure transducer placed at the inlet of the catheter. b) The strain gauges are then calibrated through static pressure. The pressure transducer values are stored together with the strain gauge voltages. Next the pressure transducer and strain gauge voltages are processed, and a curve fit is generated. Intraluminal pressure during injection is determined through this curve fit. c) catheters are finally pressurized until burst; or d) microscopic surface analysis is performed on the lumina of 1 sample of each catheter types tested.

The working principle of a strain gauge is shown in Figure 2. Deformation of the material on to which the gauges are placed results in deformation of the thin wires. This in turn results in a change in resistance which can be accurately measured. Materials deform predictably when stress is applied. Intraluminal pressure introduces such stresses in the wall of the CVCs, so strain gauges may be utilized to measure this deformation. The strain can thus be related to an intraluminal pressure when calibrated with a pressure transducer in a static closed system. Additionally, such strain gauges are typically used to determine material wear and fatigue over time. As such, they may provide information relating to such effects occurring in the CVC material with CM injection.

**Figure 2:**
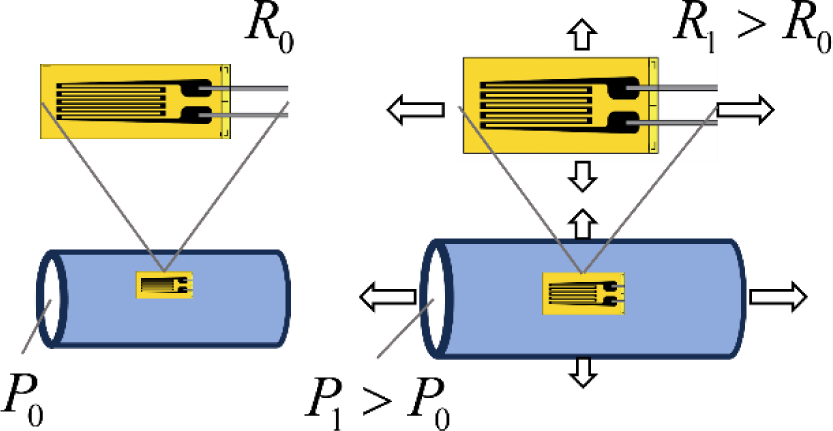
Working principle of a strain gauge. The strain gauge is placed on a surface and the resistance R is continuously measured, of which the value is R_0_ at rest with intraluminal pressure P_0_. When the material stretches, e.g. due to a higher intraluminal pressure P_1_, the strain deforms with the material. The resistance of the deformed gauge R_1_ will increase due to the increase in length and decrease in cross-sectional diameter of the wire. This can then be measured as a voltage change when amplified in a Wheatstone bridge.

A GFLAB-3-70 Low Elastic Strain Gauge (Tokyo Measuring Instruments Laboratory Co., Tokyo, Japan) was placed on the proximal side of the proximal lumen of each CVC and fastened with cyanoacrylate adhesive. As the geometry of the lumen remains more or less the same between the inlet and outlet and fluid flows from high to low pressure, pressure should be highest at this point in the CVC. The strain gauge was placed in a Wheatstone bridge to amplify the voltage change and correct the signal for effects such as temperature change. The gauges in the bridge were thus placed on a surplus CVC of the same material. When pressure was applied to the CVC, the resistance of the strain gauge changed, resulting in a change in voltage over the bridge. This voltage signal was further amplified by a CPJ Strain Gauge Conditioner (Scaime, Juvigny, France) and read in LabVIEW through a NI MyDAQ (National Instruments, Austin, TX, USA). Each tested CVC sample was fixated at two points (Figure 1a) to ensure no deformation of the catheter occurs, other than induced by pressure.

The distal lumen of each CVC was connected to a FlowSens® syringeless soft bag CM injector (Guerbet, Paris, France) with standard consumable tubing and with a PU5405 pressure transducer (ifm GmbH, Essen, Germany) fastened with a three-way stopcock just proximal to the CVC connector. The system was flushed with saline and primed so that there was CM iobitridol 350 mgI/ml (Xenetix, Guerbet, Paris, France) in the CVC prior to starting measurements. The test setup and all fluids used were operated at room temperature as strain gauges are sensitive to temperature changes. The heating element in the power injector was switched off. Because viscosity decreases with temperature which increases necessary pressures, this represents a worst-case scenario in which the fluid is not heated to body temperature prior to administration. The CM exiting the tip of the CVC was collected in a vial and reused by feeding it back into the power injector bag.

### Central venous catheters

4 samples each of 3 different types of new, unused CVCs were collected and tested:

- 13F, 250mm GamCath Dolphin Protect High Flow Double Lumen straight catheter (Baxter, Deerfield, IL, USA)
- 13F, 250mm GamCath High Flow Triple Lumen straight catheter (Baxter, Deerfield,IL, USA)
- 15.5F, 200mm Jet Medical Short-Term Free Flow pre-curved catheter (Jet Medical SA, La Chaux-de-Fonds, Switzerland)

### Flow measurement

First, 50 mL of CM was injected into each CVC at 4.5 mL/s over 11 seconds,^22^ and both the pressure transducer and strain gauge values were recorded. Due to the viscoelasticity (i.e., non-direct strain response to applying or removing pressure similar to dampening) of the material, a 5 minute resting time is needed prior to commencing the next measurement. This process was repeated 5 times. Next, 90 mL was injected at 8 mL/s over 11 seconds, corresponding to the protocol with the highest flow rate used in our center. The same 5- minute resting time was applied, for 5 measurements. The maximum pressure in the power injector was also recorded. For half the samples of each type of CVC, the protocol was reversed to 5 times 8 mL/s injection, and then 5 times 4.5 mL/s injection. Finally, the CVCs were flushed with saline.

### Pre-curved CVCs

The pre-curved CVC do not have the same “double-D” cross section, but an inner (distal) and outer (proximal) lumen (Figure 2a). A strain gauge could not be directly applied to the wall of the distal lumen. The gauges were fixed on the wall of the proximal lumen and measurements were conducted through this lumen. However, for 3 of the 4 samples, injection with the power injector was repeated with both lumina and maximum injection pressure was recorded for both to assess for similarity. The inner lumen in the remaining sample was spared for microscopic surface analysis.

### Sensor calibration and data processing

After completing the flow measurements, the tip of each catheter was clamped and the power injector was disconnected. A syringe filled with room temperature water and a screw feeder was connected to the three-way stopcock to create a closed and static system. The strain gauge was calibrated by gradually increasing the pressure to ∼800 kPa (∼120 PSI) over 2 minutes and recording the strain gauge voltage together with the known static pressure. When 120 PSI was reached, recording was stopped and pressure was released. This process was repeated 3 times for each catheter. This data was processed in MATLAB (MathWorks, Natick, Massachusetts), in which the calibration data was zero-shifted, and a 3^rd^-order polynomial was fitted. The maximum intraluminal pressure of each measurement was determined by inserting the zero-shifted maximum gauge value into the curve fit of the respective CVC. Maximum pressures were pooled per injection velocity in each CVC type. To assess material fatigue (permanent strain) and damage of the CVC material after CM injection, the resting voltages were recorded of the strain gauge after the 5-minute relaxation period of each measurement for each individual catheter. Linear regression applied was applied to these values to assess for a non-zero linear coefficient with 95% confidence.

### Burst pressure test

Three samples each of the 3 different CVC types were connected to a high-pressure manual bicycle pump with a calibrated Gems 3300 1600 kPa pressure transducer (Gems, Plainville, CT, USA) fastened through a three-way stopcock just proximal to the CVC connector. The catheter was filled with room temperature water and tip was sealed shut with a medical clamp. Pressure was manually increased until burst was recorded as a sharp decrease in the pressure transducer reading and an audible bang.

### Microscopic analysis

The CVCs that were not burst were dissected. Surface samples from the proximal and distal ends of the lumina subjected to power injection of CM were collected, sectioned and prepared for microscopy. Samples were mounted on Scanning Electron Microscopy stubs with double sides carbon stickers. Samples from the unused, proximal lumina of the same CVCs were also taken to act as control. Before imaging, all samples were sputter-coated with a layer of Gold/Palladium Images and were recorded in a GeminiSEM 300 Scanning Electron Microscope (Zeiss, Oberkochen Germany), operated at 5 kV.

## Results

### Calibration

All CVCs were successfully tested and calibrated. A 3^rd^-order polynomial curve was fitted to the calibration data of each CVC, and these curves were used to determine the intraluminal pressure during CM injection. As an example, Figure 3a shows the calibration data, curve fit and 95% confidence bounds of this fit in of a double lumen CVC tested. The complete set of calibration figures can be found in Supplemental Figures S1-S3.

**Figure 3:**
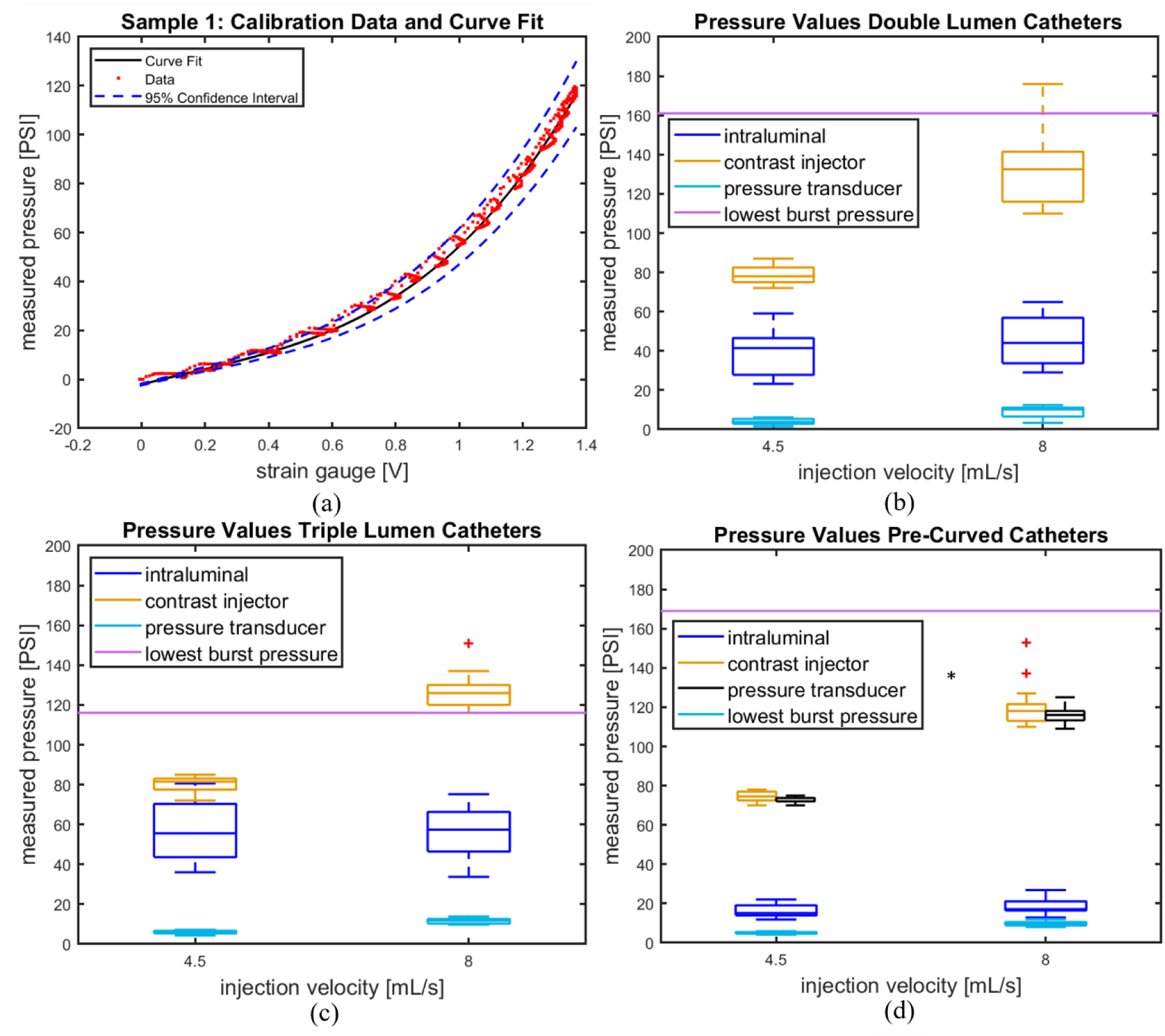
a) The calibration data, fitted 3^rd^-order polynomial and its 95% confidence bounds of catheter sample 1, used to determine intraluminal pressure with the strain gauge. b-d) Boxplots of the pump pressures, intraluminal strain gauge pressures and pressure transducer values at injection velocities 4.5 mL/s and 8 mL/s, and the burst pressures of b) the double lumen catheters; c) the triple lumen catheters; and d) the pre-curved catheters. n=4 applies to (b-d) *Only 3 samples have been tested

### Power injection of contrast medium

Figures 3b-d display the pump pressures, intraluminal strain gauge pressures (determined with the fitted calibration curves), pressure transducer values at injection velocities 4.5 mL/s and 8 mL/s, and the lowest recorded burst pressures of the 3 different CVC types.

### Material fatigue

Material fatigue, or permanent strain of the CVC, is assessed for each sample tested. Linear regression is applied to the strain gauge voltage at rest, which corresponds to the material strain at rest and fatigue. The linear regression coefficients are non-zero (p < 0.05) in every CVC tested, except CVC sample 2 (double lumen, p = 0.53). As an example, Figure 4 displays the raw strain gauge data of the first CVC measured together with the linear regression analysis performed on the resting voltages of this CVC. Figure 5 displays the linear regression analysis of CVC sample 2, the only sample in which the linear regression coefficient was not significantly non-zero. The complete set of linear regression analyses is found in Supplemental Figures S4-S6.

**Figure 4:**
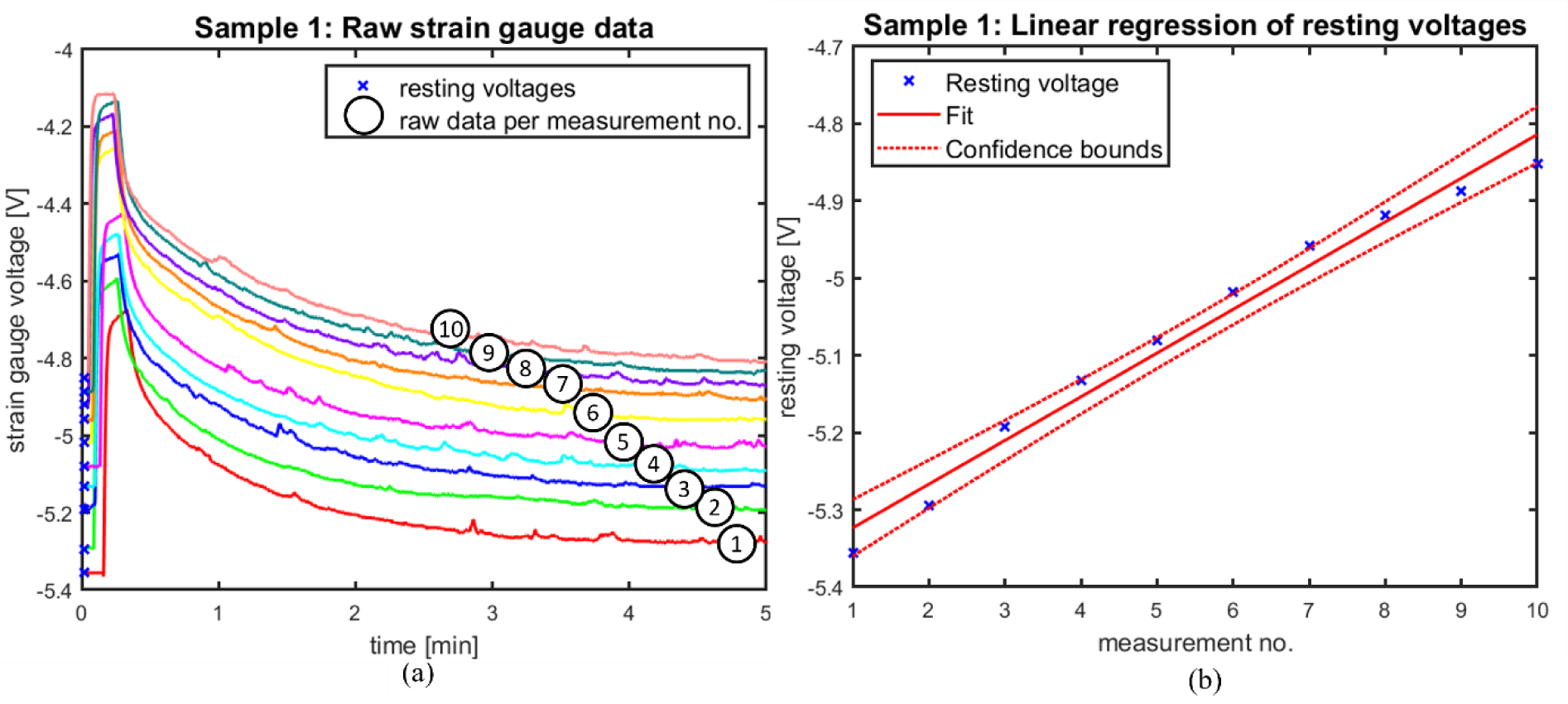
Representative regression plot of catheter 1 showing the trend in resting voltages throughout the measurements. Patterns are similar throughout the tested catheters. a) shows the raw strain gauge data over the 5-minute testing period of measurements 1 through 10 in catheter 1, as indicated by the numbers in the graph; and b) displays the regression analysis of the resting voltages as indicated in a).

**Figure 5:**
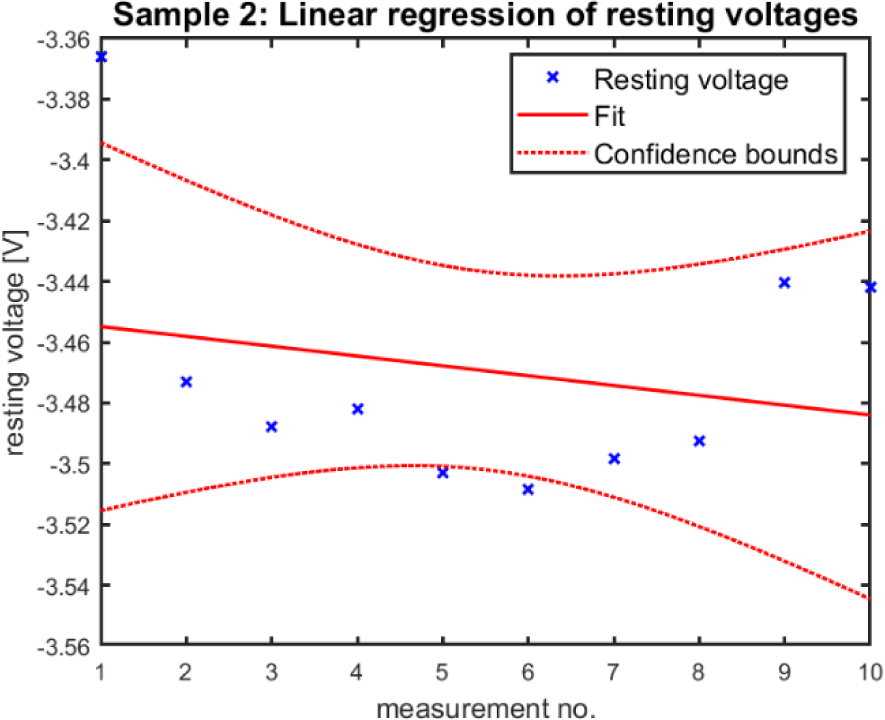
Fatigue analysis of catheter sample 2. This is the only catheter in which the linear regression coefficient was not statistically significantly non-zero.

### Burst pressure

All CVCs failed at one of the locations displayed in Figure 6 during burst pressure measurement. Burst pressures and the failure location at burst pressure are also recorded in Table 1.

**Figure 6:**
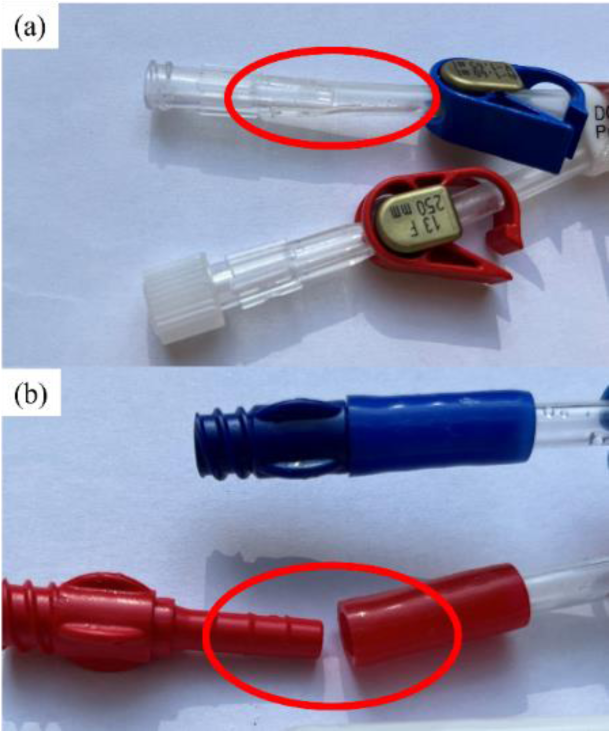
Failure modes of the central venous catheters at burst pressures: a) failure of the inlet tube resulting in rupture; and b) dislocation of the inlet tube connector.

**Table 1:** Testing regime, burst pressure and failure location for each catheter tested. Failure locations are shown in Figure 6.

| Sample no. | CVC type | Testing regime | Burst pressure [kPa]<br>([PSI]) | Failure location |
| --- | --- | --- | --- | --- |
| 1 | Double Lumen | 5x4.5mL/s; 5x8mLs | 1179 (171) | Inlet tube burst |
| 2 | Double Lumen | 5x4.5mL/s; 5x8mLs | N/A | N/A |
| 3 | Double Lumen | 5x8mLs; 5x4.5mL/s | 1158 (168) | Inlet tube burst |
| 4 | Double Lumen | 5x8mLs; 5x4.5mL/s | 1110 (161) | Inlet tube burst |
| 5 | Triple Lumen | 5x4.5mL/s; 5x8mLs | 862 (125) | Inlet tube burst |
| 6 | Triple Lumen | 5x4.5mL/s; 5x8mLs | 917 (133) | Inlet tube burst |
| 7 | Triple Lumen | 5x8mLs; 5x4.5mL/s | N/A | N/A |
| 8 | Triple Lumen | 5x8mLs; 5x4.5mL/s | 800 (116) | Inlet tube burst |
| 9 | Pre-Curved | 5x4.5mL/s; 5x8mLs | 1165 (169) | Connector dislocation |
| 10 | Pre-Curved | 5x4.5mL/s; 5x8mLs | 1248 (181) | Connector dislocation |
| 11 | Pre-Curved | 5x8mLs; 5x4.5mL/s | 1220 (177) | Connector dislocation |
| 12 | Pre-Curved | 5x8mLs; 5x4.5mL/s | N/A | N/A |

### Microscopic surface analysis

The microscopic surface analysis of the 3 CVCs examined is depicted in Figure 7. Figures 7a shows the unused lumen, and Figure 7b the proximal part of the tested lumen of the double lumen catheter. Magnification has been chosen to best illustrate the differences in the surface in each CVC. The unused and tested lumen of the triple lumen CVC are shown in Figures 7c and 7d, respectively. Finally, Figures 7e and 7f display the unused and tested lumina, respectively, of the pre-curved CVC. With the unused lumina as control, the pictures of the tested lumina show an increase in size and density of micro-cracks in the double and triple lumen CVCs. These micro-cracks present themselves as clear black lines in the microscopy images. However, this contrast is less apparent in the pre-curved CVC.

**Figure 7:**
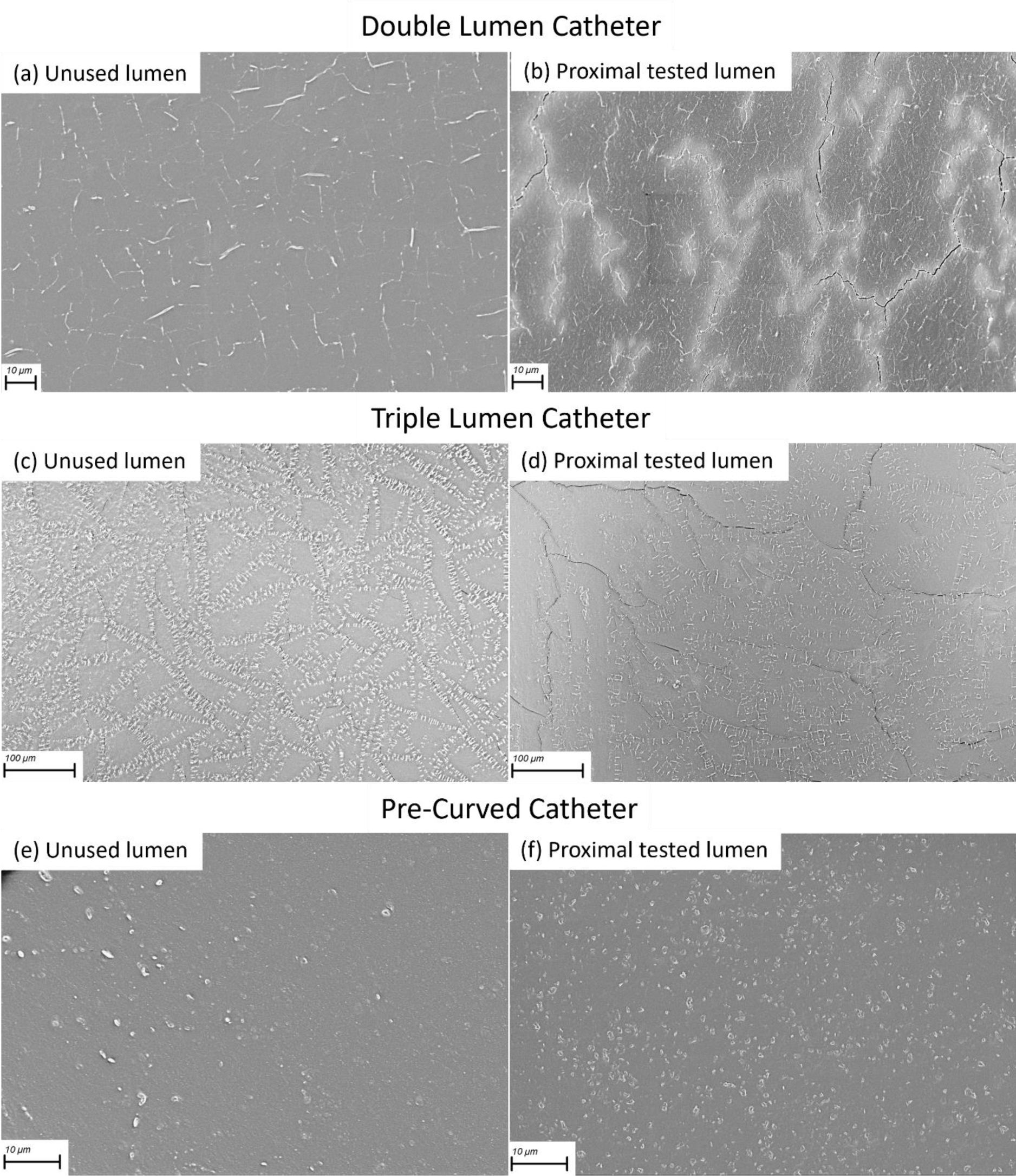
Surface analysis through scanning electron microscopy of: a) unused lumen, and b) proximal part of the tested lumen, of the double lumen catheter; c) unused lumen, and d) proximal part of the tested lumen of the triple lumen catheter; e) unused lumen, and f) proximal part of the tested lumen, of the pre-curved catheter. Small white dots and lines visible in all images are identified as pores in the material. The black lines are indicative of micro-cracks. There are notable differences in the luminal surfaces across the various types of catheters. The magnifications used were selected to best illustrate the surface changes in each catheter type before and after exposure to contrast medium injection.

## Discussion

In this study it was observed that intraluminal pressures during power injection of CM remain well below burst pressure. By using strain gauges and a pressure transducer in the flow circuit, the hypothesis could be supported that strain gauges provide a more accurate measurement of intraluminal pressure. Moreover, the strain gauge measurements suggest material fatigue and damage can occur to the CVCs with repeated use. Surface analysis of the CVCs was performed to assess microscopic damage to the material, in which a greater incidence of micro-cracks was noticed after testing. This data can guide healthcare professionals in responsible use of CVCs for power injection of CM.

The measured pressure data is in line with other experiments in literature in showing that the intraluminal pressure always remained sufficiently low.^3–8^ Bursting pressures are also comparable.^3, 8^ The CM injector pressure was higher than the intraluminal pressure, which is necessary to overcome the frictional losses in the tubing and CVCs. A pressure gradient is required for a fluid to flow. However, the pump pressure also remained below the bursting pressure in instances with 4.5 mL/s injection, and most instances with 8mL/s. The relatively large variation in injector pressure, with several outliers may be explained by the pump exerting an increased pressure on the fluid bag when almost empty, which was a constant trend. The intraluminal pressures also show a large variation in values. This may for a large part be explained by the sensitivity of the gauges to environmental factors, measurement errors and the additional step of calibrating the values with a fitted curve. Regardless, they always remained below the burst pressure and pump pressure.

The intraluminal pressures in the different types of CVCs show some discrepancy. Although pump pressures were quite similar throughout the measurements, the lumina of the types of catheters have dissimilar cross-sections. Due to the shape and larger diameter of the pre-curved CVCs, the tested lumen in this CVC type has the greatest cross-sectional area, which in the measurements corresponds to the lowest intraluminal pressures. Contrarily, the highest intraluminal pressures were found in the triple lumen CVCs, which have the smallest cross-sectional area as a result of the extra lumen. Despite the geometrical differences between the lumina of the pre-curved CVCs, the pump pressures were similar through both when injecting at the same velocity. This suggests intraluminal pressures are also similar through each lumen.

Although the CM injector exerted a significantly greater pressure for an 8 mL/s injection compared to 4.5 mL/s, the intraluminal pressures in the strain gauges appear very similar. An explanation could lie in the frictional losses of the fluid flowing through a substantial length of tubing before entering the CVC. Frictional losses are greater with a higher velocity, and due to the small diameter of the tubing and high viscosity of the CM, it can be expected that the losses contribute considerably to pressure losses. Additionally, the three-way stopcock present in the flow circuit may have introduced more turbulence in the flow which could have resulted in a slight drop in pressure and affected the measurements downstream.

The laws of fluid mechanics state that the pressure transducer should measure a greater pressure than the strain gauges, as it is placed further upstream. Therefore, it is an interesting finding that the pressure measured by the pressure transducer during the injections is noticeably lower than the intraluminal pressure as measured by the strain gauges. An explanation may lie in geometrical conditions. Pressure transducers placed into a flow circuit inevitably interfere with the flow itself, as most often, a three-way connection is placed to connect the transducer that introduces dead space in which fluid is static. These stagnant zones act as reservoirs of pressure, exerting localized effects that can distort the overall pressure profile in such locations. Turbulent eddies may form in regions of abrupt flow constriction or expansion, causing fluctuations in pressure and velocity. In systems with rapidly changing flow rates, the presence of dead space can thus impede the propagation of pressure waves and dampen fluid oscillations, leading to distorted pressure transducer readings.^23^ This dead space will thus not always show a direct pressure response to fluidic flow, which impedes accuracy of the measurement. These phenomena may contribute to the rather large differences in measured pressures throughout previous in vitro studies (2-70 PSI at 4.5 mL/s velocities with similar catheters). ^3, 4, 7, 9^ As the strain gauges do not interfere with the flow and they measure pressure directly at the location of interest, the pressures they measured are more likely to be valid.

Repetitive CM injections revealed a gradual increase in strain throughout the measurements. The fatigue analysis of the strain values implies that material damage accumulates throughout the repeated measurements. Consistency in peak amplitudes suggests that the adhesive interface between the strain gauge and CVC remained intact. In all CVCs, except one, this coefficient is statistically significant. In CVC sample 2 the non-significant coefficient appears to result from the last 2 measurements having a far higher resting voltage. This likely occurred through accidental interference with the CVC during testing. Moving the CVC can result in small changes in deformation which are also registered due to the high sensitivity of strain gauges.

The fatigue measured in the CVCs over the power injections recorded by the strain gauges is further supported by the microscopic surface analysis. The microscopy shows an increase in size and density of micro-cracks between the unused and tested luminal surfaces in the double and triple lumen CVCs. These cracks seem to propagate along what appear to be small pores in the material. Propagation is likely induced by the pressure of the CM injection. Electron microscopy in literature of similar dialysis CVCs exposed to normal use and explanted from patients do not show such clear micro-cracks.^24, 25^ This upholds the notion that the high pressure and viscosity of the CM can cause permanent damage to the CVC surfaces. In the pre-curved CVCs, no such phenomenon is clearly visible. This could be due to the intraluminal pressures in the pre-curved CVCs being lower, but the material itself also appears smoother.

As the CVC material appears to stretch after CM injection, permanent stretching and offset may have occurred prior to calibration. The validity of calibration data can thus be questioned. Although most regression coefficients are significantly non-zero, the actual changes in resting voltage are relatively small compared to the height of the voltage peaks. Moreover, the heights of the voltage peaks (maximum value minus resting voltage) remain fairly constant throughout the 5 measurements at the different injection velocities in all CVCs. The effects on the determined intraluminal pressure values are therefore expected to be limited.

These data suggest that intraluminal pressure values during CM injection are sufficiently low as to not cause rupture of the CVCs included in our study. Failure of the CVCs at bursting pressure always occurred at the inlet, which in clinical use will remain outside the body. However, the stretching of the material and increase in micro-crack size and density suggested damage accumulates with repeated CM injection. The micro-cracks may increase risks of thrombus formation in the CVC but remains to be studied. With the velocities tested, incidental use likely carries little risk, especially with relatively new CVCs that have not yet been frequently subjected to intraluminal pressure. However, these medical devices remain uncertified for this use case. Therefore, caution must be taken and caretakes must understand that liability is shifted away from the manufacturer when using such CVCs for CM injection. Clinical use of CVCs for this purpose has previously been shown to be safe and effective, provided adequate protocols are in place.^16^ For example, a sufficiently safe maximum pressure should be placed on the injector. If a blockage would be present in the CVC preventing flow, the injector pressure would equate to the intraluminal pressure in the CVC, which could cause damage or even rupture.

This study has several limitations. Firstly, intraluminal pressures were measured in an in vitro setup, in which conditions present in the human body are not fully simulated. The CVCs were not maintained at body temperature. Testing at body temperature could have increased flexibility of the material to some extent. Contrarily, fibrous tissue tends to form on the exterior of the catheters when placed in the body, which can increase rigidity and therefore intraluminal pressure. Venous pressure at the tip was not present, which may have increased intraluminal pressure. However, venous pressure is typically far lower than the pressures measured in the system. Moreover, intraluminal pressure is only measured at one location in the catheters. Although pressure close to the inlet is expected to be highest, it does not fully examine the pressure and stress profile throughout the entire CVC and therefore does not consider any stress or pressure concentrations that could occur. Additionally, the CM was at a lower temperature than typically used when injecting into the body. The viscosity will therefore have been higher, and the resulting pressures likely overestimated. Any changes in fluid composition due to evaporation during reuse of the fluid are not considered. Moreover, the fatigue analysis suggests that stretching of and damage to the material occurs after repeated use. However, it is not known how the material fatigue relates to the strength of the material, and to which extent safe use may be maintained. This should receive attention in future studies. Finally, the CVCs are not subjected to normal clinical handling. Human factors like bending, pushing and pulling during placement may have a significant effect on the material integrity.

## Conclusion

This study aimed equip healthcare professionals with the knowledge for responsible administration of CM injection for angiography through CVCs. Healthcare professionals must understand that liability shifts away from the manufacturer if such a device is not certified for this use case. Strain gauges allowed intraluminal pressure measurements at the location of interest as well as an estimation of stretch that the intraluminal pressure induces. Intraluminal pressures during CM injection remained below bursting pressure in all instances, but permanent stretch of the material was recorded. Microscopic surface analysis revealed an increase in micro-crack size and density in the material after repeated use with high intraluminal pressures. These findings suggest that incidental use of CVCs for CM injection does not directly cause issues, but damage may accumulate if CM injections are done repetitively. Caution must therefore always be taken, especially after repeated use.

## Data Availability

All data referred to in the manuscript is available upon reasonable request.

## Acknowledgments

The authors would like to express their gratitude to J. Brenkman of the TU Delft for assisting in the assembly of the strain gauge test setup. They also thank R.I. de Koning of the LUMC for his help with the electron microscopy.

## Supplemental Materials

Figures S1-S6

## List of abbreviations

CM: contrast medium/media
CVC: central venous catheter

## References

1. Contrast Media Safety Committee. ESUR Guidelines on Contrast Agents v10.0 [Internet]. European Society of Urogenital Radiology. 2018. 0–45 p. Available from: http://www.esur.org/fileadmin/content/2019/ESUR_Guidelines_10.0_Final_Version.pdf

2. Davenport, Matthew S. Daniella AJC e. al. ACR Manual On Contrast Media 2020 ACR Committee on Drugs and Contrast Media. Stanford Protocol. 2023. 131 p.

3. Beckingham T, Roberts A, St. John A, O’Callaghan G. Bursting pressure of triple-lumen central venous catheters under static and dynamic loads. J Vasc Access [Internet]. 2017;18(5):430–5. Available from: 10.5301/jva.5000776

4. Macha DB, Nelson RC, Howle LE, Hollingsworth JW, Schindera ST. Central venous catheter integrity during mechanical power injection of iodinated contrast medium. Radiology. 2009;253(3):870–8.

5. Zamos DT, Emch TM, Patton HA, D’Amico FJ, Bansal SK. Injection Rate Threshold of Triple-Lumen Central Venous Catheters:. An In Vitro Study. Acad Radiol. 2007;14(5):574–8.

6. Herts R, Cohen MAH, Zepp C, Einstein M. Power Injection of Intravenous Contrast Material through Central Venous Catheters for CT: In Vitro Evaluation. Radiology. 1996;200(3):731–5.

7. Hollander S, Mojibian H, Emery M, Tal MG. Power injection of iodinated intravenous contrast material through acute and chronic hemodialysis catheters. J Vasc Access. 2013;13(1):61–4.

8. Smirk C, Soosay Raj T, Smith AL, Morris S. Neonatal percutaneous central venous lines: fit to burst. Arch Dis Child - Fetal Neonatal Ed. 2009 Jul 1;94(4):F298–300.

9. Schwartz FR, Lewis DS, King AE, Murphy FG, Howle LE, Kim CY, et al. Hemodialysis catheter integrity during mechanical power injection of iodinated contrast medium for computed tomography angiography. Abdom Radiol. 2021 Jun 1;46(6):2961–7.

10. Coyle D, Bloomgarden D, Beres R, Patel S, Sane S, Hurst E. Power Injection of Contrast Media via Peripherally Inserted Central Catheters for CT. J Vasc Interv Radiol. 2004 Aug 1;15(8):809–14.

11. Donnelly LF, Dickerson J, Racadio JM. Is hand injection of central venous catheters for contrast-enhanced CT safe in children? AJR Am J Roentgenol. 2007 Dec;189(6):1530–2.

12. Buijs SB, Barentsz MW, Smits MLJ, Gratama JWC, Spronk PE. Systematic review of the safety and efficacy of contrast injection via venous catheters for contrast-enhanced computed tomography. Eur J Radiol Open. 2017 Jan 1;4:118–22.

13. The European Commission. Regulation (EU) 2017/745 of The European Parliament and of the Council on medical devices. 2017.

14. Code of Federal Regulations. Title 21 Food and Drugs Chapter I - Food and Drug Administration Department of Health and Human Services Subchapter H - Medical Devices Part 801 —Labeling Subpart A—General Labeling Provisions. Vol. 30. 2014.

15. Plumb AAO, Murphy G. The use of central venous catheters for intravenous contrast injection for CT examinations. Br J Radiol. 2011 Mar 1;84(999):197–203.

16. Herts BR, O’Malley CM, Wirth SL, Lieber ML, Pohlman B. Power injection of contrast media using central venous catheters: Feasibility, safety, and efficacy. Am J Roentgenol. 2001;176(2):447–53.

17. Sanelli PC, Deshmukh M, Ougorets I, Caiati R, Heier LA. Safety and feasibility of using a central venous catheter for rapid contrast injection rates. AJR Am J Roentgenol [Internet]. 2004 [cited 2024 May 6];183(6):1829–34. Available from: https://pubmed.ncbi.nlm.nih.gov/15547237/

18. Dodd JD, Kalva S, Pena A, Bamberg F, Shapiro MD, Abbara S, et al. Emergency cardiac CT for suspected acute coronary syndrome: Qualitative and quantitative assessment of coronary, pulmonary, and aortic image quality. Am J Roentgenol. 2008 Sep 23;191(3):870–7.

19. Stein PD, Fowler SE, Goodman LR, Gottschalk A, Hales CA, Hull RD, et al. Multidetector Computed Tomography for Acute Pulmonary Embolism. N Engl J Med. 2006;354.

20. Anderson DR, Kahn SR, Rodger MA, Kovacs MJ, Morris T, Hirsch A, et al. Computed Tomographic Pulmonary Angiography vs Ventilation-Perfusion Lung Scanning in Patients With Suspected Pulmonary Embolism: A Randomized Controlled Trial. JAMA. 2007 Dec 19;298(23):2743–53.

21. Radiological Society of the Netherlands. Safe use of contrast media - part 2 [Internet]. 2019 [cited 2024 Jul 22]. Available from: www.radiologen.nl

22. Beckingham T, Roberts A, St. John A, O’Callaghan G. Bursting pressure of triple-lumen central venous catheters under static and dynamic loads. J Vasc Access. 2017;18(5):430–5.

23. White F, Ng C, Saimek S. Fluid Mechanics (7th ed.). 7th ed. New York City, NY: McGraw-Hill Education; 2011.

24. Kanaa M, Wright MJ, Sandoe JAT. Examination of tunnelled haemodialysis catheters using scanning electron microscopy. Clin Microbiol Infect. 2010;16:780–6.

25. Lucas TC, Tessarolo F, Veniero P, Caola I, Piccoli F, Haase A, et al. Hemodialysis catheter thrombi: Visualization and quantification of microstructures and cellular composition. J Vasc Access [Internet]. 2013 Sep 4 [cited 2024 Jul 9];14(3):257–63. Available from: https://journals.sagepub.com/doi/10.5301/jva.5000142?url_ver=Z39.88-2003&rfr_id=ori%3Arid%3Acrossref.org&rfr_dat=cr_pub++0pubmed

